# Ethnic and regional variation in hospital mortality from COVID-19 in Brazil

**DOI:** 10.1101/2020.05.19.20107094

**Authors:** Pedro Baqui, Ioana Bica, Valerio Marra, Ari Ercole, Mihaela van der Schaar

**Affiliations:** Núcleo de Astrofísica e Cosmologia (Cosmo-ufes), Universidade Federal do Espírito Santo, 29075-910, Vitória, ES, Brazil; Department of Engineering Science, University of Oxford, Parks Road, Oxford OX1 3PJ, UK; The Alan Turing Institute, 96 Euston Rd, London NW1 2DB, UK; PPGCosmo & Departamento de Física, Universidade Federal do Espírito Santo, 29075-910, Vitória, ES, Brazil; University of Cambridge Department of Medicine, Addenbrooke’s Hospital, Hills Road, Cambridge CB2 0QQ, UK; Centre for Mathematical Sciences, University of Cambridge, Wilberforce Rd, Cambridge CB3 0WA, UK; Department of Electrical Engineering, University of California, Los Angeles, Los Angeles, CA 90095 USA

## Abstract

**Background:** The COVID-19 pandemic is quickly spreading throughout Brazil, which is rapidly ascending the ranking of countries with the highest number of cases and deaths. A particularly unstable federal regime and fragile socioeconomic situation is likely to have contributed to the impact of the disease. Amid this crisis there is substantial concern in the possible socioeconomic, geopolitical and ethnic inequity of the impact of COVID-19 on the country’s particularly diverse population.

**Methods:** We performed a cross-sectional observational study of COVID-19 hospital mortality using observational data from the SIVEP-Gripe dataset. We present descriptive statistics to quantify the COVID-19 pandemic in Brazil. We assess the importance of regional factors such as education, income and health either on a state-by-state basis or by splitting Brazil into a North and a Central-South region. Mixed-effects survival analysis was used to estimate the effects of ethnicity and comorbidity at an individual level in the context of regional variation.

**Findings:** Our results show that, compared to *branco* comparators, hospitalised *pardo* and *preto* Brazilians have significantly higher risk of mortality, with hazard ratios and 95% CI of 1.47 (1.33-1.58) and 1.32 (1.15-1.52), respectively. In particular, *pardo* ethnicity was the second most important risk factor (after age). We also found that hospitalised Brazilians in North regions tend to have more comorbidities than in the Central-South, with similar proportions between the various ethnic groups. Finally, we found that states in the North have a higher hazard ratio as compared to the Central-South, and that Rio de Janeiro obtained one of the highest hazard ratios, similar to the ones of the more underdeveloped Pernambuco and Amazonas.

**Interpretation:** Our results can be interpreted according to the interplay of two independent, but correlated, effects: i) mortality by COVID-19 increases going North (vertical effect), ii) mortality increases for the *pardo* and *preto* population (horizontal effect). We speculate that the vertical effect is driven by increasing levels of comorbidity in Northern regions where levels of socioeconomic development are lower, whereas the horizontal effect may be related to lower levels of healthcare access or availability (including intensive care) for *pardo* and *preto* Brazilians. For most states the vertical and horizontal effects are correlated giving a larger cumulative mortality. However, Rio de Janeiro was found to be an outlier to this trend: It has an ethnicity composition (horizontal effect) similar to the states in the North region, despite high levels of socioeconomic development (vertical effect). Our analysis motivates an urgent effort on the part of Brazilian authorities to consider how the national response to COVID-19 can better protect *pardo* and *preto* Brazilians as well as the population of poorer states from their higher death risk from SARS-CoV-2 infection.

**Funding:** None.

## INTRODUCTION

The COVID-19 pandemic has created an unprecedented worldwide strain on healthcare. Whilst early reports from East Asia and Europe meant that Brazil was well positioned to implement non-pharmaceutical interventions, Brazilians, like those in many developing countries, have limited access to testing and social security. The former makes it difficult to assess the growth of the pandemic, while the latter prevents a sizable fraction of society from engaging in physical distancing. This has been further complicated by an unstable federal government^1^ that has failed to support measures such as social distancing and attempted to downplay the gravity of the pandemic. Worryingly, as of May 19^th^, Brazil ranks fourth worldwide for total COVID-19 cases and sixth for deaths, with the highest estimated rate of transmission in the world (*R*_0_ = 2.81),^2^ second only to USA for the daily increase of confirmed cases and deaths.

Worldwide there is substantial interest in the emerging societal inequity of the impact of COVID-19, and there is emerging evidence to suggest variability in the impact of the disease across ethnicities in a variety of settings including in the UK,^3-4^ USA^6-8^ and Norway.^9^ Brazil’s population is particularly diverse, comprising many races and ethnic groups. The Brazilian Institute of Geography and Statistics (IBGE) racially classifies the Brazilian population in five categories (percentages as of 2010): *branca* (47.7%), *parda* (43.1%), *preta* (7.6%), *amarela* (1.1%) and *indígena* (0.4%). We will use the Portuguese terms throughout this work. This IBGE classification is based on color and, as in international practice, individuals are asked to self identify as either: *Branco* (‘white’), *preto* (‘black’), *amarelo* (‘yellow’), *indígeno* (‘Amerindians’) or *pardo*. The term *pardo* is particularly complex one and is used in Brazil to refer to people of mixed ethnic ancestries: *pardo* Brazilians represent a diverse range of ethnic backgrounds. While *branco* and *pardo* Brazilians together comprise the majority of the population, with approximately equal proportions, their distribution varies considerably regionally. For example, the population in the South macro-region is 78% *branca* and 17% *parda*, while the North macro-region’s population is 23% *branca* and 67% *parda*.^10^

Research in context

Evidence before this study

Brazil is a highly ethnically and socioeconomically diverse country. The severe impact of COVID-19, coupled with an unstable federal regime, may make it particularly susceptible to outcome inequities. Although the issue of the disproportionate effect of COVID-19 on ethnic groups has been debated in the Brazilian media, quantitative / systematic studies assessing the ethnic and regional variation in mortality from SARS-CoV-2 in Brazil are lacking. Added value of this study

We found that hospitalised *pardo* and *pret.o* Brazilians have statistically significant higher mortality compared to a *branco* comparator group. In particular, *pardo* ethnicity was the second most important risk factor after age. We also found that mortality by COVID-19 increases in socioeconomically comparable Northern regions, and that Rio de Janeiro has an exceptionally high risk as compared to its neighboring states.

Implications of all the available evidence

Our results have serious social implications: *pardo* and *preto* Brazilians have, on average, less economic security, are less likely to be able to stay at home and work remotely, and comprise a significant proportion of health and care workers. We hope that this analysis assists the authorities in better directing and aligning their response to COVID-19 in order to protect *pardo* and *preto* Brazilians from their higher death risk from SARS-CoV-2. Our results also indicate that the states in the North and Northeast macroregions are more vulnerable to the COVID-19 pandemic, an issue that merits further urgent attention by the federal government.

The combination of the severity of the outbreak, governmental failure to implement non-pharmaceutical interventions and complex social and ethnic societal composition makes Brazil a particularly important as well as interesting country in which to study the impact of COVID-19. In this work, we analyze COVID-19 hospital mortality from the prospectively collected SIVEP-Gripe (“Sistema de Informação da Vigilância Epidemiológica da Gripe”) respiratory infection registry data, which is maintained by the Ministry of Health for the purposes of recording cases of Severe Acute Respiratory Syndrome (SARS) across both public and private hospitals. Using this rich dataset, we characterize the COVID-19 pandemic in Brazil, particularly with regard to risk factors related to comorbidities, symptoms and ethnicity similarly to previous analyses in countries such as the UK.^5,11-13^

## METHODS

Our analysis is based on the SIVEP-Gripe public dataset.^14^ As of the time of access, this contains epidemiological data for 99,557 patients from different states. Each entry has 139 features, including symptoms, age, sex, ethnicity and comorbidities. Applying the condition of having tested positive for COVID-19 leaves data for 19,940 patients. As we are interested in the relation between ethnicity and health risk, we then consider only data with ethnicity recorded, leaving data for 12,221 patients. Furthermore, we consider only the subset that was hospitalized: this is our base dataset of 11,321 patients, see Fig. 1. The date of diagnosis spans the time interval from 27^th^ February 2020 to 4^th^ May 2020. Our analysis employs descriptive statistics to quantify the COVID-19 pandemic in Brazil, and Cox regression to estimate hazard ratios. Cox regression needs a reliable date of outcome, and, therefore, we further restrict the dataset as depicted in Fig. 1.

**Figure 1.**
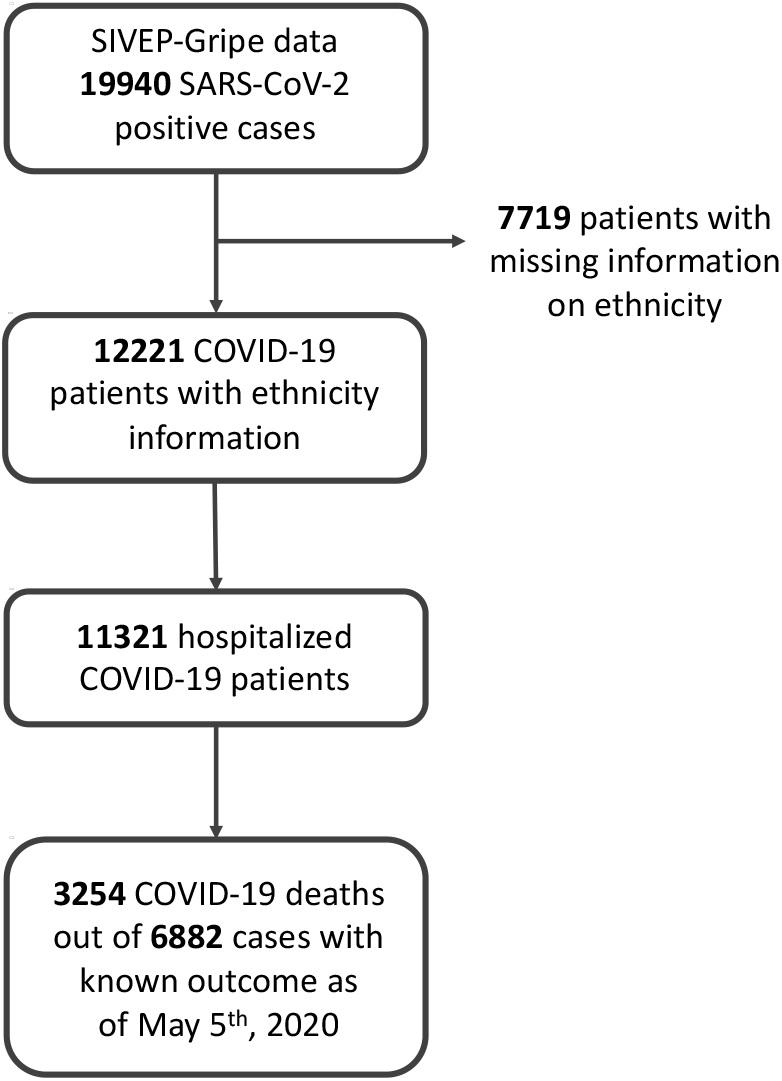
STROBE flowchart of SIVEP-Gripe data used in this study.

Brazil is a very heterogeneous country, a federation composed of the union of 26 states and a federal district. In the Cox analysis we assess the importance of factors such as education and income of each Brazilian state, as also health indexes such as the number of ICU beds, ventilators, doctors and nurses in the public and private heath care system.^15^

Brazil is divided geopolitically into 5 macroregions:

- North: Acre (AC), Amapá (AP), Amazonas (AM), Pará (PA), Rondônia (RO), Roraima (RR), Tocantins (TO);
- Northeast: Alagoas (AL), Bahia (BA), Ceará (CE), Maranhão (MA), Paraíba (PB), Pernambuco (PE), Piauí (PI), Rio Grande do Norte (RN), Sergipe (SE);
- Central-West: Distrito Federal (DF), Goiás (GO), Mato Grosso (MT), Mato Grosso do Sul (MS);
- Southeast: Espírito Santo (ES), Minas Gerais (MG), Rio de Janeiro (RJ), São Paulo (SP);
- South: Paraná (PR), Rio Grande do Sul (RS), Santa Catarina (SC).

For descriptive purposes we chose to dichotomise the data into two maximally contrasting regions based on similar education (literacy, higher education and school drop-out rates), income (per-capita gross domestic product, salary and poverty level) and health (life expectancy, child mortality and food security). Ethnicity was not considered at this point. We do not consider a finer subdivision in order to maximize statistical significance. The two regions that we consider are:

- “Central-South” which comprises the Central-West, Southeast and South macroregions for a total of 9278 patients (81%)
- “North” which comprises the North and Northeast macroregions for a total of 2043 patients (19%).

As shown in the supplementary materials, this division appears quite naturally once the socioeconomic factors are combined and is also the customary one when one splits Brazil into a Northern and a Southern region.

The SIVEP-Gripe file that is used at the hospitals includes information on comorbidities and symptoms. We interpret missing values as absence of comorbidities or symptoms as we assume that doctors mainly filled in the information for the present comorbidities. Missing values are also present for ICU admissions. In this case we consider missing values as non-admissions to ICU. The number of patients with missing data for symptoms and comorbidities can be found in the supplementary materials, as per STROBE requirement.

To investigate the effects of record-level risk factors, we fitted a multivariate mixed-effects Cox regression model to estimate hazard ratios for in-hospital mortality. We used patient-level clinical features, namely age group, sex, ethnic group and comorbidities as fixed effects, with geographical location (i.e. state of Brazil) as the random effect, similarly to previous analyses in the context of UK NHS hospitals.^16^ For the categorical variables age group and ethnic group, we used Age < 40 and *branco* as reference categories, respectively. Statistical testing was used to evaluate the proportional hazards assumption^17^ and we did not find any statistically significant evidence that it was violated (p=0.11).

## RESULTS

### Descriptive statistics

Fig. 2 shows the distribution of COVID-19 cases among the Brazilian states. One sees that SP, RJ and AM have the highest number of cases, both absolute and per capita.

**Figure 2.**
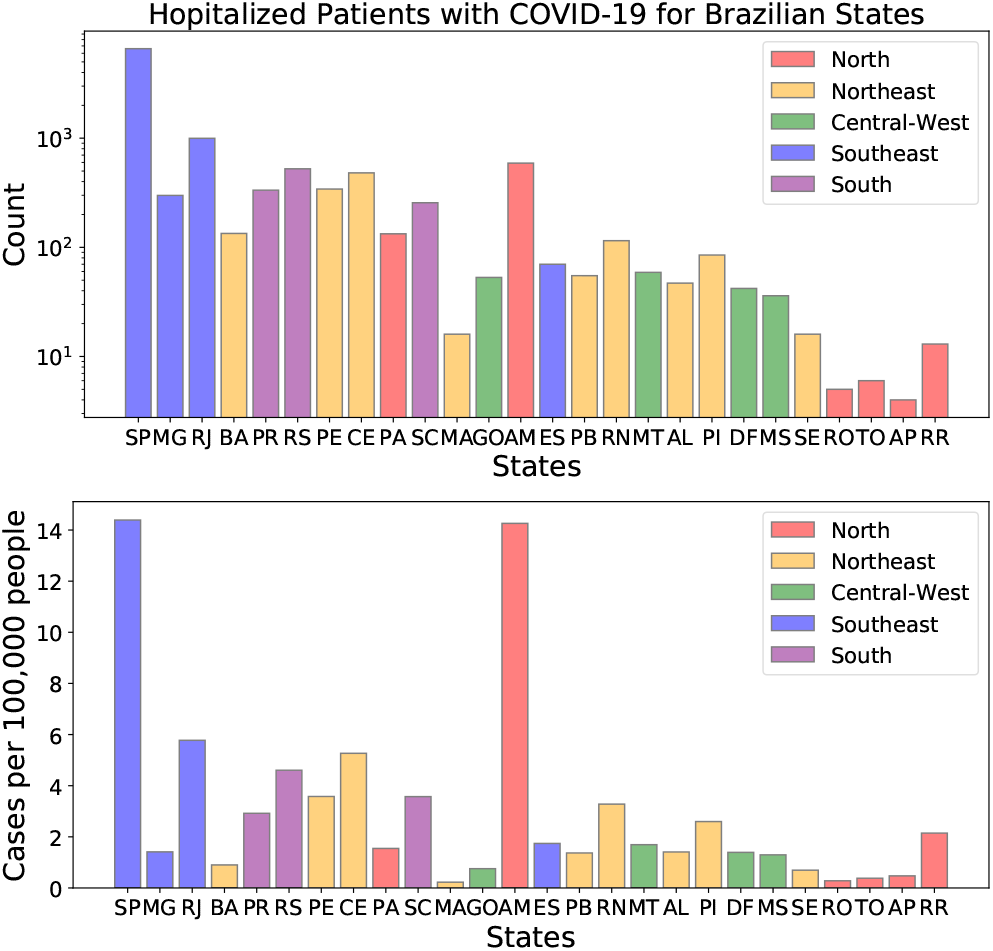
Distribution of the dataset considered in this work (11321 patients) among the Brazilian states. States are ordered according to their population, larger on the left.

Table I shows the distribution of demographic characteristics and comorbidities among survivors and nonsurvivors of COVID-19, while Table II shows the ethnic composition of patients at each stage of the COVID-19 trajectory.

**Table I.** Demographic characteristics and coexisting conditions among survivors and non-survivors of COVID-19.

|  | survivors | non-survivors |
| --- | --- | --- |
| North (2043 data) | 479 (35.5%) | 871 (64.5%) |
| Age [yr] | 46.9 ± 19.3 | 65.3 ± 16.0 |
| Female sex [no. (%)] | 218 (39.3%) | 337 (60.7%) |
| Male sex [no. (%)] | 261 (32.8%) | 534 (67.2%) |
| Ethnic group [no. (%)] |  |  |
| *branca* | 89 (39.6%) | 136 (60.4%) |
| *parda* | 366 (34.9%) | 683 (65.1%) |
| *preta* | 16 (31.4%) | 35 (68.6%) |
| *amarela* | 5 (29.4%) | 12 (70.6%) |
| *indígena* | 3 (37.5%) | 5 (62.5%) |
| Comorbidities [no. (%)] |  |  |
| Cardiovascular disease | 95 (22.6%) | 325 (77.4%) |
| Asthma | 22 (62.9%) | 13 (37.1%) |
| Diabetes | 74 (19.9%) | 297 (80.1%) |
| Pulmonary disease | 15 (29.4%) | 36 (70.6%) |
| Obesity | 13 (22.4%) | 45 (77.6%) |
| Immunosuppression | 28 (57.1%) | 21 (42.9%) |
| Renal disease | 13 (18.8%) | 56 (81.2%) |
| Liver disease | 4 (23.5%) | 13 (76.5%) |
| Neurological disease | 7 (21.2%) | 26 (78.8%) |
| Central-South (9278 data) | 3564 (59.2%) | 2457 (40.8%) |
| Age [yr] | 52.2 ± 16.6 | 67.0 ± 15.8 |
| Female sex [no. (%)] | 1525 (60.4%) | 1001 (39.6%) |
| Male sex [no. (%)] | 2039 (58.3%) | 1456 (41.7%) |
| Ethnic group [no. (%)] |  |  |
| *branca* | 2548 (62.0%) | 1560 (38.0%) |
| *parda* | 728 (53.7%) | 627 (46.3%) |
| *preta* | 215 (50.6%) | 210 (49.4%) |
| *amarela* | 69 (54.8%) | 57 (45.2%) |
| *indígena* | 4 (57.1%) | 3 (42.9%) |
| Comorbidities [no. (%)] |  |  |
| Cardiovascular disease | 936 (44.9%) | 1147 (55.1%) |
| Asthma | 158 (64.8%) | 86 (35.2%) |
| Diabetes | 641 (42.1%) | 880 (57.9%) |
| Pulmonary disease | 115 (34.2%) | 221 (65.8%) |
| Obesity | 130 (48.9%) | 136 (51.1%) |
| Immunosuppression | 104 (40.0%) | 156 (60.0%) |
| Renal disease | 87 (27.2%) | 233 (72.8%) |
| Liver disease | 25 (37.9%) | 41 (62.1%) |
| Neurological disease | 84 (29.7%) | 199 (70.3%) |

From Table I one sees that, in both the North and Central-South regions, survivors are younger and more likely to be white and female, while non-survivors are more likely to be *preto* and *pardo*, respectively (results regarding the other ethnicities are more difficult to interpret due to the lower numbers). In the North region, non-survivors display a great prevalence of almost all comorbidities. This suggests that the overall health condition in the North region is worse compared to the Central-South region. This is confirmed by the significantly larger percentage of non-survivors in the North.

The same trend is observed by comparing the total number of hospitalized patients to the number of deaths (fifth column of Table II). Again we see that mortality is significantly higher in the North as compared to the Central-South and that *preto* and *pardo* Brazilians are less likely to survive the virus. In other words, we are observing two independent effects: i) mortality by COVID-19 increases going North (vertical effect), ii) mortality increases for the *preto* and *pardo* population (horizontal effect).

To better assess the prevalence of comorbidities, Fig. 3 shows the distributions by ethnicity and number of comorbidities, for survivors and non-survivors, excluding *indígeno* patients because of the small numbers. There is a notable North/Central-South asymmetry with more non-survivors in the former.

**Figure 3.**
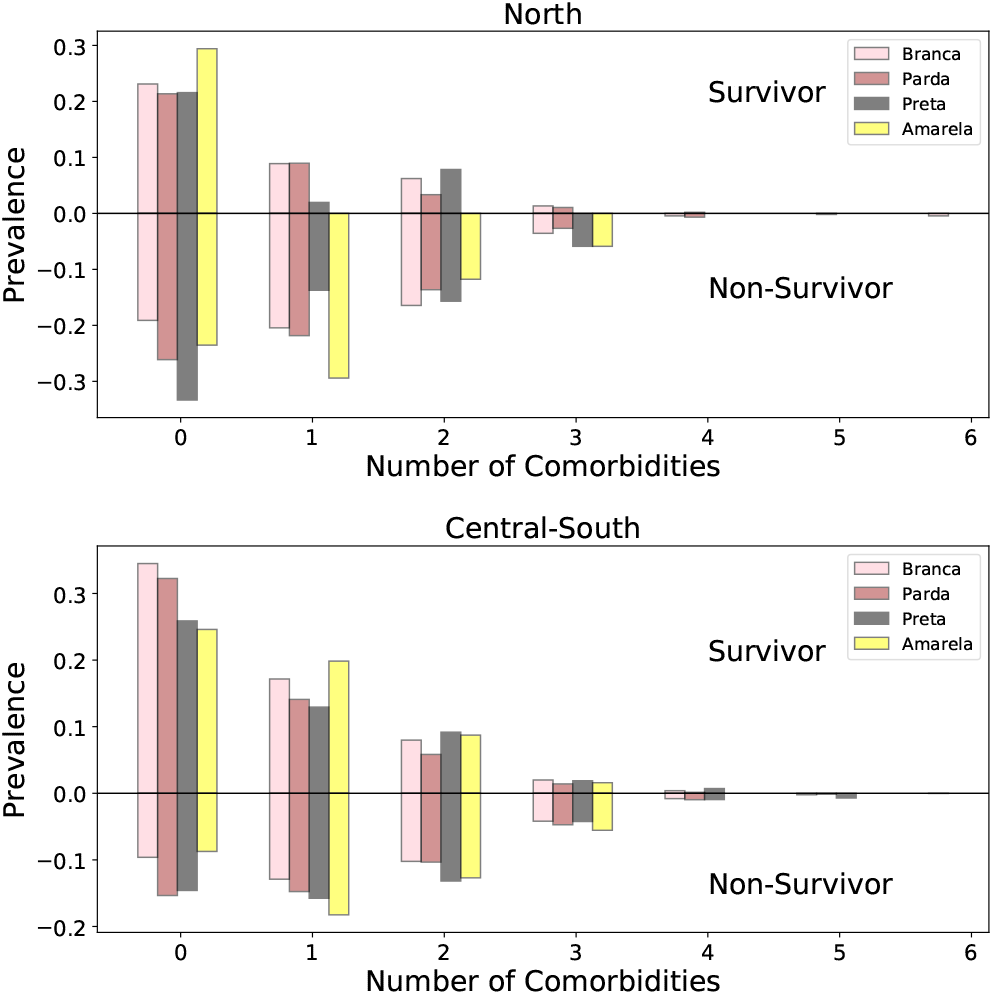
Distributions according to number of comorbidities and ethnicity. The normalization is such that all the fractions of a given ethnicity add to unity. We exclude *indígeno* patients for clarity due to their small numbers.

Fig. 4 shows the distributions according to number of symptoms and ethnicity, split according to mortality. We consider the following symptoms: fever, cough, sore throat, shortness of breath, respiratory discomfort, SpO2 < 95%, diarrhea, and vomiting. Most patients present between 3 and 6 symptoms, suggesting again that in this dataset, it is the more severe presentations that were tested for COVID-19.

**Figure 4.**
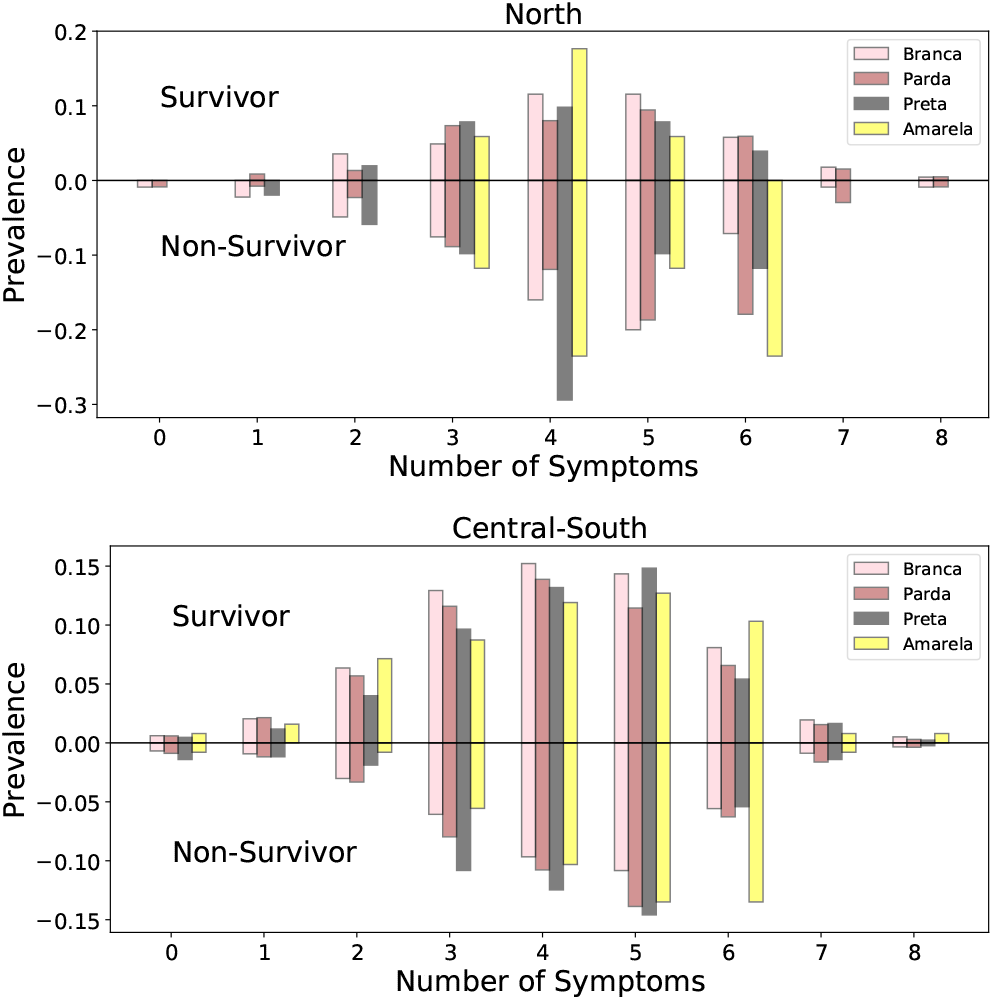
Distributions according to number of symptoms and ethnicity. The normalization is such that all the fractions of a given ethnicity add to unity. We exclude *indígeno* patients for clarity due to their small numbers.

Fig. 5 presents the distributions according to age and ethnicity for survivors and non-survivors. The expected trend that younger patients are more likely to survive is much more pronounced in the North. Also, younger *pardo* and *preto* Brazilians seem to be less likely to survive the virus compared to *branco* Brazilians, with the difference being more pronounced in the Central-South region.

**Figure 5.**
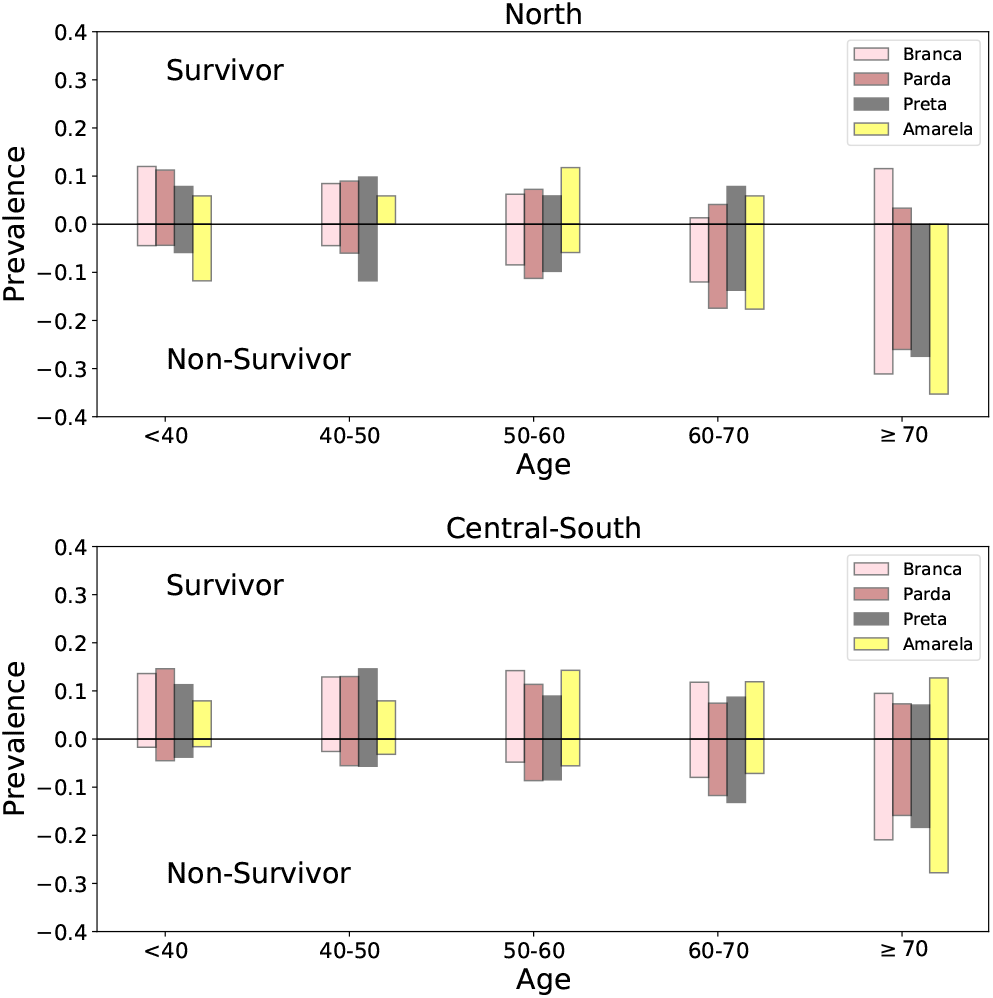
Distributions according to age and ethnicity. The normalization is such that all the fractions of a given ethnicity add to unity. We exclude *indígeno* patients for clarity due to their small numbers.

Fig. 6 shows hazard ratios with 95% confidence intervals for all clinical features (fixed effects) considered in the fitted multivariate mixed-effects Cox model for inhospital mortality. Compared to *branco* comparators, hospitalised *pardo* and *preto* Brazilians have significantly higher risk of mortality. In particular, *pardo* ethnicity was the second most important risk factor (after age).

**Figure 6.**
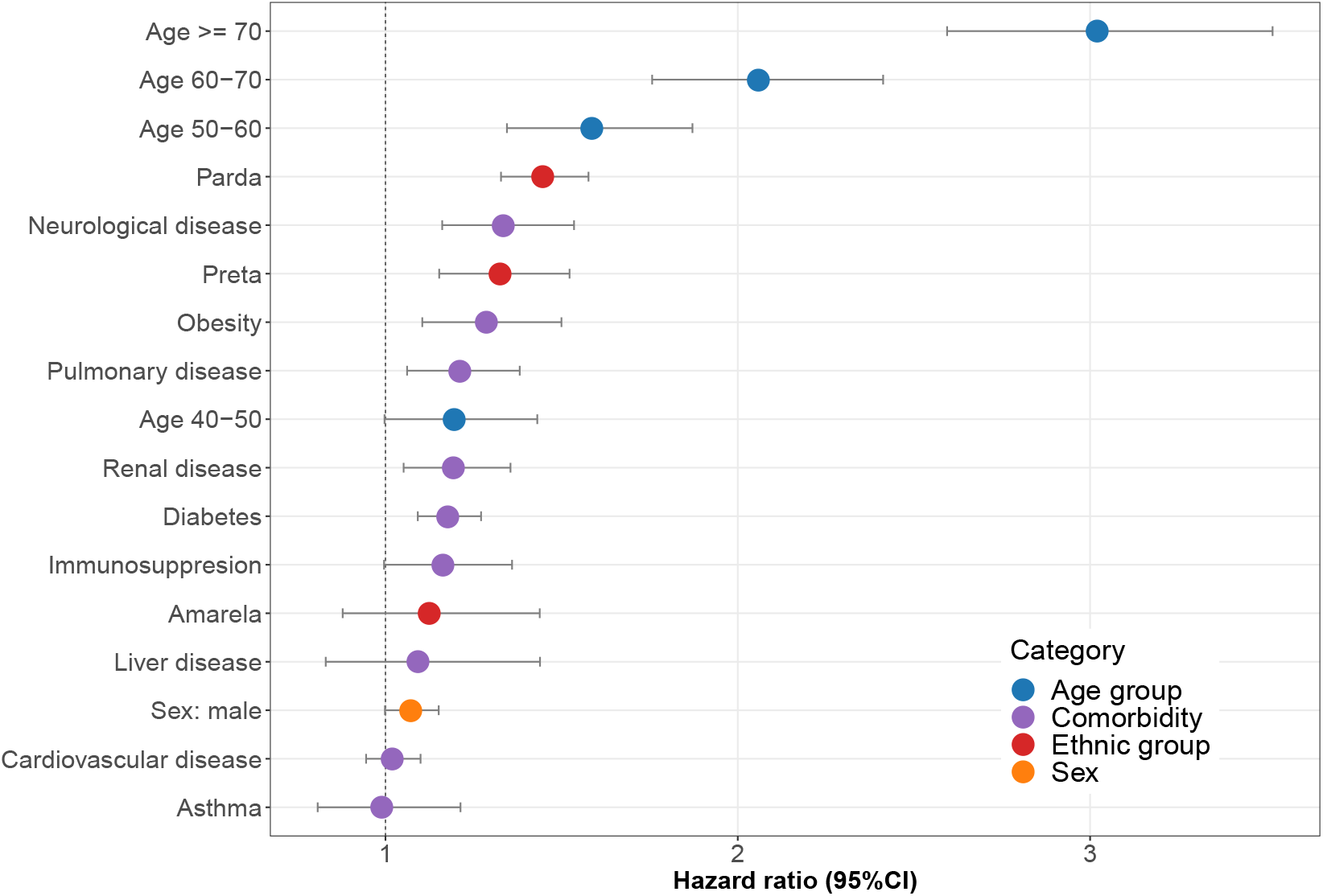
Fixed effects (hazard ratio) estimates with their 95% confidence intervals for in-hospital mortality.

Fig. 7 shows hazard ratios with 95% confidence intervals for all different states in Brazil (random effects) considered in the fitted multivariate mixed-effects Cox model for in-hospital mortality. A substantial between-states variation is apparent. The states in the North region tend to have higher hazard ratios than the ones belonging to the Central-South region, further justifying our approach of splitting Brazil into two sets. This also agrees with the significantly larger percentage of non-survivors in the North as shown in Tables I and II (vertical effect discussed earlier).

**Table II.** Ethnic composition of patients at each stage of the COVID-19 trajectory.

| North<br>(2043 data) | Hospitalization | ICU<br>Admission | Death | Death | Death | Death |
| --- | --- | --- | --- | --- | --- | --- |
|  |  |  |  | Hospitalization | (not ICU) | (ICU) |
| *branca* | 342 (16.7%) | 127 (19.4%) | 136 (15.6%) | 39.8% | 69 (14.9%) | 67 (16.5%) |
| *parda* | 1567 (76.7%) | 481 (73.8%) | 683 (78.4%) | 43.6% | 368 (79.3%) | 315 (77.4%) |
| *preta* | 85 (4.2%) | 26 (4.0%) | 35 (4.0%) | 41.2% | 20 (4.3 %) | 15 (3.7%) |
| *amarela* | 36 (1.8%) | 13 (2.0%) | 12 (1.4%) | 33.3% | 5 (1.1 %) | 7 (1.7%) |
| *indígena* | 13 (0.6%) | 5 (0.8%) | 5 (0.6%) | 38.5% | 2 (0.4%) | 3 (0.7%) |
| Central-South<br>(9278 data) | Hospitalization | ICU<br>Admission | Death | Death | Death | Death |
|  |  |  |  | Hospitalization | (not ICU) | (ICU) |
| *branca* | 6291 (67.8%) | 2344 (69.4%) | 1560 (63.5%) | 24.8% | 616 (60.3%) | 944 (65.8%) |
| *parda* | 2112 (22.8%) | 731 (21.6%) | 627 (25.5%) | 29.7% | 278 (27.2%) | 349 (24.3%) |
| *preta* | 667 (7.2%) | 220 (6.5%) | 210 (8.6%) | 31.5% | 108 (10.6%) | 102 (7.1%) |
| *amarela* | 195 (2.1%) | 82 (2.4%) | 57 (2.3%) | 29.2% | 19 (1.9%) | 38 (2.7%) |
| *indígena* | 13 (0.1%) | 2 (0.1%) | 3 (0.1%) | 23.1% | 1 (0.1%) | 2 (0.1%) |

**Figure 7.**
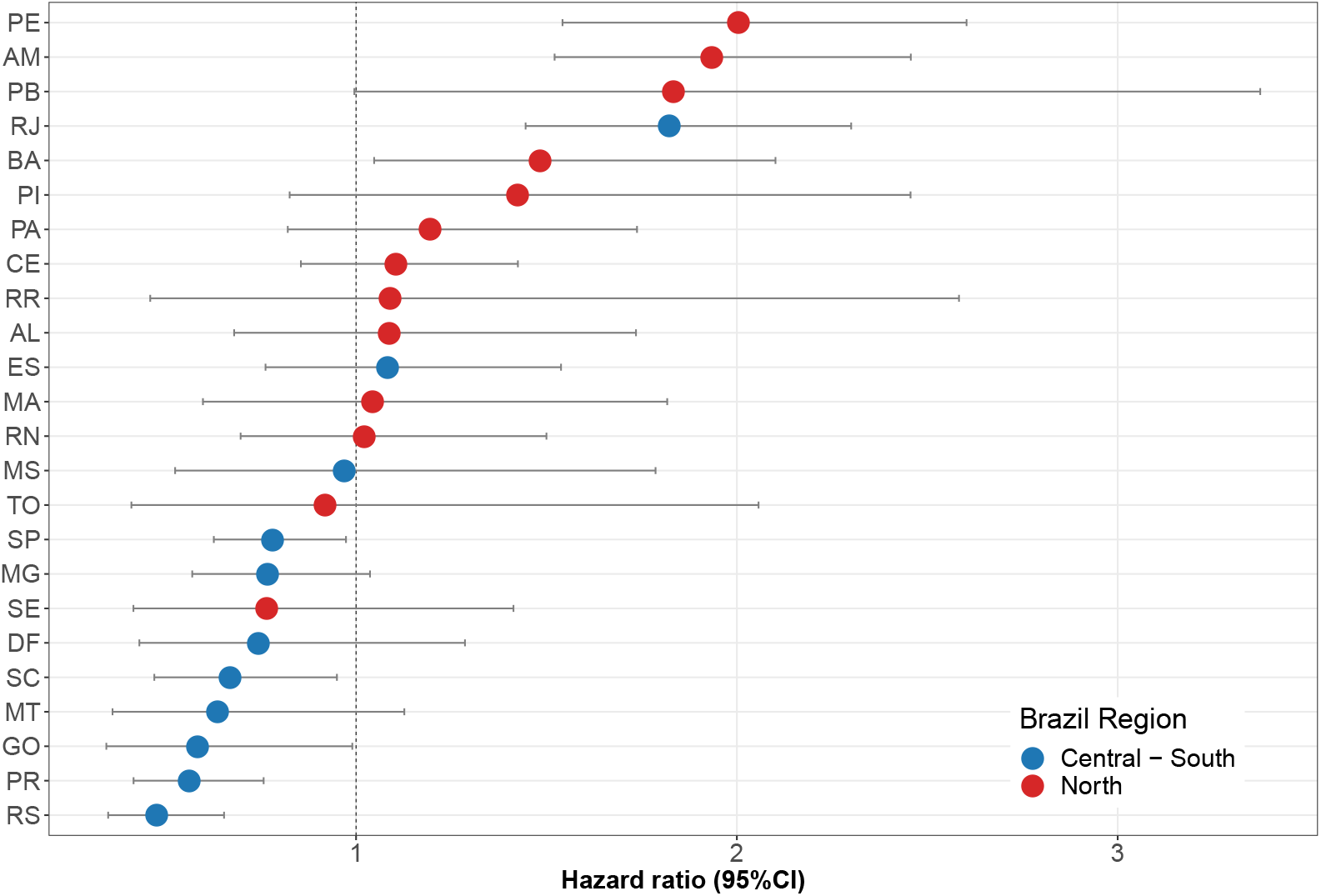
Random effects (hazard ratio) showing between-Brazilian-state variation with their 95% confidence intervals.

## DISCUSSION

We present, to our knowledge, the largest study of COVID-19 hospital survival in Brazil. We show that survivors are younger^18^ with female preponderance^19^ and tend to have fewer comorbidities,^20^ replicating worldwide findings. However, we also report a number of other important sociodemographic trends which are specific to Brazil.

There is significant regional variation in both case-mix and outcome. The high number of cases (both in absolute and per-capita terms) seen in SP, RJ and AM (Fig. 2) are interesting. Of note, these regions are important ports of entrance to Brazil: AM hosts the Free Economic Zone of Manaus and most of the international flights route through SP and RJ: in 2019 7.7 million international passengers landed in SP and 2.2 million in RJ^21^ (further details in the supplementary materials). Therefore, these states are characterized by a higher international circulation of people. Furthermore, both SP and RJ are characterized by a particularly high population density and the COVID-19 outbreak coincided with the rainy season in AM, which is usually associated with a peak in respiratory infections.

In order to explore regional differences we split the Brazilian states into socioeconomically comparable North and Central-South regions. The finding of a higher comorbidity burden in hospitalised patients in the North region is concordant with a lower life expectancy^22^ and is also borne out by differences in the average age of survivors/non-survivors between the North and Central-South regions, as well as the significantly larger percentage of non-survivors in the North. In addition, we found that, in both regions, survivors are more likely to be *branco* and that *branco* Brazilians are more likely to be admitted to ICU than *pardo* Brazilians. In other words, the increased death rate of *pardo* Brazilians seems to be due in part to non-ICU admission, raising concerns regarding the organization of public and private medical resources.

It is also noteworthy that the distribution of hospitalized COVID-19 patients between the Central-South (81%) and North (19%) regions in our data is discordant with the actual population sizes of these two regions (64% and 36% respectively).^10^ This highlights the diversity in the country and may, at least in part, be due to either intrinsically lower hospitalization rates in the North region and/or disproportionate impact of COVID-19 in populous areas such as São Paulo and Rio de Janeiro (both in the Central-South region), the populations of which total about 20 million people.

The disparity in comorbidity among hospitalized patients between the North and Central-South regions is also striking (Table I), with non-survivors displaying a greater prevalence of almost all comorbidities in the North: Only asthma and immunosuppression are below hospital prevalence 70%. By contrast, for the Central-South region, the only comorbidities affecting more than 70% of non-survivors are renal and neurological diseases. Whilst a number of structural explanations are of course possible, this is likely to reflect poorer health status overall in the North region which, again, has a substantially larger percentage of non-survivors.

The disproportionately large percentage of comorbidity-free survivors in the Central-South region (Fig. 3) is remarkable. We may speculate that this may be due to differences in comorbidity ascertainment either due to structural differences in the way data is collected - perhaps comorbidity data was less available from patients who were sicker at the time of presentation - or because less severe patients, perhaps with concerns regarding their comorbid health, presented to hospital preferentially in the Central-South region. It is interesting to observe that *branco* and *pardo* Brazilians have a similar number of comorbidities in these populations. Therefore, it seems that comorbidities are not associated with ethnicity in the group studied, but are correlated with regional socioeconomic development (education, income and health).

However, an interplay between ethnic and regional socioeconomic factors is apparent in Fig. 5 with younger *pardo* and *preto* Brazilians seemingly less likely to survive the disease compared to the *branco* population; the difference being more pronounced in the Central-South region. It is worth placing this in the context of the typical life-expectancy in Brazil of 76 years (as of 2017).^23^ By way of comparison the life expectancy in the European Union is 80.9 years.^24^ Note also that the average life expectancy varies significantly by region, being higher in the Central-South than in the North: Santa Catarina has a life expectancy of 79.4 years while that of Maranhão is 70.9. This provides a baseline for the trend shown in Fig. 5.

From the ethnic distribution of patients admitted to ICU shown in Table II it appears that *branco* Brazilians are more likely than *pardo* Brazilians to be admitted to ICU once hospitalised. Also, by comparing percentages of hospitalization with deaths, one sees that *branco* Brazilians are more likely to survive than *pardo* Brazilians. However, by comparing total hospitalization with deaths after ICU admission, one sees more similar proportions between both ethnicities.

It appears that there are substantial ethnic differences in the proportion of patients admitted to ICU, and this also varies between North and Central-South regions. The greater proportion of deaths without admission to ICU for *pardo* is noteworth. This is likely to reflect higher levels of access to private healthcare for *branco* as compared to *pardo* Brazilians as ICU admission policies are known to differ between public and private hospital settings.^25^ This may suggest that the lower rate of admission to ICU may be a factor for the increased proportion of deaths among *pardo* Brazilians. Note that the distribution of comorbidities, symptoms and age does not show strong ethnic variations, especially between *pardo* and *branco*. That the proportions of the different ethnicities admitted to ICU with COVID-19 are similar to those in the full 2019 SIVEP-Gripe dataset^14^ in Table III suggests that this is not a specific feature of COVID-19.

**Table III.** Ethnic composition of 32,338 patients hospitalized and then admitted to ICU according to the 2019 SIVEP-Gripe dataset.

| Brazil<br>(32773 data) | Hospitalization | ICU |
| --- | --- | --- |
|  |  | Admission |
| *branca* | 18723 (57.9%) | 7314 (60.9%) |
| *parda* | 11772 (36.4%) | 3983 (33.2%) |
| *preta* | 1379 (4.3%) | 539 (4.5%) |
| *amarela* | 234 (0.7%) | 99 (0.8%) |
| *indígena* | 230 (0.7%) | 72 (0.6%) |

At an individual level survival analysis showed that, after age, the most important factor for hospital mortality was being of *pardo* or, to a lesser extent, *preto* ethnicity with hazard ratios of 1.47(1.33-1.58) and 1.32(1.15-1.52) (95% CI) compared to the *branco* baseline. The other risk factors largely replicate worldwide findings although we see that male sex is perhaps slightly less of a risk factor compared to what has been found in other series.^19^ This ethnic inequity has important social roots and implications: *pardo* and *preto* Brazilians have, on average, less economic security, are less likely to be able to stay at home and work remotely, and comprise a significant proportion of health and care workers, making them disproportionately the most vulnerable to COVID-19.^26^ We also performed the Cox regression separately on the patients in the North and South regions, on patients in São Paulo and also excluding the São Paulo and Amazonas states (the ones with the highest number of cases per capita, see Fig. 2) to test robustness of our findings to outliers: The results were qualitatively consistent and did not change our conclusions that ethnicity is a key risk factor (details in the supplementary materials).

Examining the magnitude of the random effect estimates, we see substantial variation in hazard by region. The states that belong to the North region tend to have higher hazard ratios than the ones belonging to the Central-South region, which provides additional justification for splitting Brazil into two socioeconomically similar sets. It is also in agreement with the significantly larger percentage of non-survivors in the North as shown in Table I. Incorporating number of ICU beds/ventilators and nurses per 100 million inhabitants for each region as proxies for physical availability of healthcare resources did not qualitatively change our result (supplementary materials) suggesting a more fundamental difference in healthcare access and trajectory.

Rio de Janeiro, despite high standards of education, income and health, has one of the highest hazard ratios, very similar to those seen for the less developed Pernambuco and Amazonas regions. It is possible that this represents the effect of a highly populous area on viral spread. At the same time, Table IV shows the ethnic distribution for the seven states with highest number of cases, together with their hazard ratios. States in the North region have a hazard ratio greater than unity and a high proportion of *pardo* Brazilians, while states in the Central-South region have a hazard ratio less than one and a higher proportion of *branco* Brazilians. The notable exception is Rio de Janeiro, which exhibits a much higher hazard ratio than the neighboring state of São Paulo and an ethnicity similar to the states in the North region.

**Table IV.** Distribution of ethnic groups (percentages) for the Brazilian states with highest number of COVID-19 cases (ordered by number of cases, see Fig. 2) together with their hazard ratio with 95% CI. Red marks states in the North region and blue in the Central-South region, similarly to Fig. 7.

| State | *branca* | *pardo* | *preto* | *amarela* | *indígena* | Hazard ratio |
| --- | --- | --- | --- | --- | --- | --- |
| SP | 63.7 | 29.4 | 5.4 | 1.4 | 0.1 | 0.78(0.63–0.97) |
| RJ | 47.4 | 39.6 | 12.1 | 0.8 | 0.1 | 1.82(1.45–2.30) |
| AM | 21.2 | 69 | 4.1 | 0.9 | 4.8 | 1.93(1.52–2.46) |
| RS | 83.2 | 10.6 | 5.5 | 0.3 | 0.3 | 0.47(0.35–0.65) |
| CE | 31.6 | 62.3 | 4.6 | 1.2 | 0.2 | 1.10(0.85–1.42) |
| PE | 36.5 | 55.5 | 6.4 | 1 | 0.6 | 2.00(1.54–2.60) |
| PR | 70.1 | 25.4 | 3.1 | 1.2 | 0.2 | 0.56(0.41–0.76) |

Many *preto* Brazilians may identify themselves as *pardo* Brazilians.^27^ For this reason, it is reasonable to consider the *preto* and *pardo* populations together. Indeed, as seen from our analysis, both ethnic groups share higher percentages of non-survivors and higher hazard ratios.

The results of our analysis can then be interpreted according to the interplay of two independent, but correlated, effects:

a. mortality by COVID-19 increases going North: ‘vertical’ effect,
b. mortality increases for the *pardo* and *preto* population: ‘horizontal’ effect.

We may speculate that the vertical effect is due to expected increasing levels of comorbidities (or poorly controlled comorbidities) and general healthcare access, which we might expect in regions such as the North where socioeconomic levels are lower. We similarly postulate that the horizontal effect is likely driven by the greater reliance on publicly-funded healthcare in *pardo* and *preto* communities whereas *branco* Brazilians on average have greater access to private healthcare. This may explain the lower ICU admission for *pardo* as compared to *branco* Brazilians as ICU admission policy and availability vary substantially between public and private hospitals.^25^

For most states the vertical and horizontal effects are correlated, giving a larger cumulative mortality. Indeed, lower socioeconomic development correlates with a larger *pardo* and *preto* population. Again, Rio de Janeiro is an outlier with an ethnic composition (horizontal effect) that is similar to the states in the North region, but high levels of development (vertical effect) more akin to those of Central-South states. In other words, Rio de Janeiro violates the typical correlation between vertical and horizontal effects seen elsewhere.

Whilst we believe our work is the most comprehensive of its kind to date in Brazil, there are a number of limitations which are worthy of discussion. Limitations and possible biases in case ascertainment cannot be ruled out, in common with all observational / database research. Ethnicity is missing in 39% of our data. This is not specific to COVID-19 (32% for the full SIVEP-Gripe dataset) but we cannot be sure that this is not subject to bias. We have limited our analysis to patients who were hospitalised since testing in the community is more likely to be biased according to local factors. However, again here, we cannot be sure that the availability of testing practice is homogeneous even in this population. Indeed, the fact that a large fraction of patients that have tested positive are admitted to the hospital clearly shows that testing, at least as far as this dataset is concerned, is performed only when symptoms are severe, indicating in turn that the number of COVID-19 cases in Brazil is likely to be much higher than suggested by available data.^28^^29^

It is possible that health-seeking behaviour varies with both ethnicity and region and late presentation may be an important determinant of ultimate hospital outcome. We are not able to consider this in our analysis as physiological severity at hospital presentation / admission data is not available in the SIVEP-Gripe dataset. However, a recent UK study did not demonstrate an important effect of physiological severity^16^ at least for ICU mortality, suggesting a high degree of homogeneity at admission, at least for this group. We cannot comment on whether this applies to Brazil or for the hospital population as a whole, and this would be an interesting area for future research.

We conducted our study during the Brazilian outbreak in the hope of generating actionable results. However, the analysis of early data introduces the possibility of leadtime and outcome ascertainment bias, although since our data encompasses a period substantially longer than the typical hospital stay, we hope that this does not affect our results qualitatively in any substantial way.

Whilst we have focused on hospital mortality, it is important to appreciate that we do not have data on out-of-hospital mortality (which may be appreciable) and neither can we robustly address the question of access to hospital services both by region or ethnicity / socioeconomic status. As such, a consideration of hospital mortality is likely to substantially underestimate the true effects of COVID-19 and it is plausible to assume that healthcare availability inequities would be further amplified in patients who are not hospitalized.

In conclusion, we present evidence suggesting a higher risk of death among *pardo* and *preto* Brazilians and in the North region. We hope that this analysis assists the authorities in better directing and aligning their response to COVID-19 in order to protect *pardo* and *preto* Brazilians from their higher death risk from SARS-CoV-2. Our results also indicate that the states in the North and Northeast macroregions are more vulnerable to the COVID-19 pandemic, an issue that merits further urgent attention by the federal government.

## Data Availability

SIVEP-Gripe data is publicly available from the Ministry of Health website.

http://plataforma.saude.gov.br/coronavirus/dados-abertos/sivep-gripe/

## CONTRIBUTORS

MvdS conceived the research question. All authors designed the study and analysis plan. VM obtained the epidemiological and socioeconomic data. PB carried out the analysis with descriptive statistics. IB carried out the analysis with Cox regression. VM drafted the initial version of the manuscript. AE oversaw the clincal review of the methods and manuscript. All authors critically reviewed early and final versions of the manuscript.

## DATA SHARING

SIVEP-Gripe data is publicly available from http://plataforma.saude.gov.br/coronavirus/dados-abertos/sivep-gripe. Our analysis code is made available in the supplementary material.

## ACKNOWLEDGMENTS

We would like to thank Roberto Andre Kraenkel for help with the SIVEP-Gripe catalog, and the CCE/UFES COVID-19 group and Fabiana V. Campos for useful comments and discussions.

